# A rapid, cost efficient and simple method to identify current SARS-CoV-2 variants of concern by Sanger sequencing part of the spike protein gene

**DOI:** 10.1101/2021.03.27.21252266

**Authors:** Tue Sparholt Jørgensen, Kai Blin, Franziska Kuntke, Henrik K. Salling, Thomas Y Michaelsen, Mads Albertsen, Helene Larsen

## Abstract

In 2020, the novel coronavirus, SARS-CoV-2, caused a pandemic, which is still raging at the time of writing this. Many countries have set up high throughput RT-qPCR based diagnostics for people with COVID-19 symptoms and for the wider population. In addition, with the use of whole genome sequencing (WGS) new lineages of SARS-CoV-2 have been identified that have been associated with increased transmissibility or altered vaccine efficacy, so-called Variants of Concern (VoC). WGS is generally too labor intensive and expensive to be applied to all positive samples from the diagnostic tests, and often has a turnaround time too long to enable VoC focused contact tracing. Here, we propose to use Sanger sequencing for the detection of common variants of concern and key mutations in early 2021, using a single set of the recognized ARTIC Network primers. The proposed setup relies entirely on materials and methods already in use in diagnostic RT-qPCR labs and on existing infrastructure from companies that have specialized in cheap and rapid turnaround Sanger sequencing. In addition, we provide an automated mutation calling software (https://github.com/kblin/covid-spike-classification). We have validated the setup on 195 SARS-CoV-2 positive samples, and we were able to profile >85% of RT-qPCR positive samples, where the last 15% largely stem from samples with low viral count. At approximately 4€ per sample in material cost, with minimal hands-on time, little data handling, and a turnaround time of less than 30 hours, the setup is simple enough to be implemented in any SARS-CoV-2 RT-qPCR diagnostic lab. Our protocol provides results that can be used to focus contact-tracing efforts and it is cheap enough for the tracking and surveillance of all positive samples for emerging variants such as B.1.1.7, B.1.351 and P.1 as of January 2021.

## Introduction

The pandemic caused by the severe acute respiratory syndrome coronavirus 2 (SARS-CoV-2) is a crisis faced by nearly every country in the world at the time of writing this article. Throughout 2020, the spread of SARS-CoV-2 caused millions of deaths, and more than 100 million people have been diagnosed with COVID-19, primarily by the use of Real-Time quantitative Polymerase Chain Reaction (RT-qPCR) diagnostic tests. In the RT-qPCR tests, the relative number of viral copies in a sample is estimated by the threshold cycle (Ct) value, where a higher value indicates fewer viral copies. Variants of the virus are identified by whole genome sequencing (WGS) based mutation calls and compared to the originally published genome^1^ or by variant specific qPCR tests^2,3^. Numerous protocols for SARS-CoV-2 WGS exist, with different versions of the ARTIC Network protocols being the most commonly used^4^. WGS derived genomes are deposited in the GISAID database^5^, where researchers can then use them to analyze outbreaks. Tracking new mutations throughout the population has led to the identification of several VoCs of SARS-CoV-2, such as the lineages B.1.1.7^6^, B.1.351^7^, and P.1^8^. The lineage B.1.1.7 carries the spike protein N501Y mutation, and is thought to be more transmissive^9^, and this mutation is also found in the B.1.351 and P.1 variants. The lineages B.1.351 and P.1 in addition are characterized by a E484K mutation, which has been reported to significantly reduce antibody binding^10^. This could potentially lower the efficacy of current vaccines. Monitoring VoCs makes it possible to tailor the societal response for maximal containment of SARS-CoV-2 and at the lowest cost. While WGS is crucial for surveillance work, it is not optimally suited for near real-time contact tracing of VoCs, as it is more expensive, requires a centralized setup and a high degree of technical and bioinformatics expertise. Instead, we have developed a cost efficient, rapid, and simple method for SARS-CoV-2 variant mutation typing using no additional equipment to that used for the diagnostic RT-qPCR SARS-CoV-2 test. (Figure 1). At a cost of approx. 4€ per reaction in materials, and approximately one and a half hours of hands on time per 96 samples (Figure 1), this system can readily be implemented for large scale typing of all positive samples, without the use of specialized equipment. The material cost is several times cheaper than the cost of WGS, regardless of the sequencing platform used^4^. Our method consists of an RT-PCR with a single ARTIC Network primer set, using the same enzyme mix used in diagnostic RT-qPCR tests, and using Sanger sequencing to sequence a 1001bp amplicon in the spike protein. Sanger sequencing as a technology is more than 40 years old^11^, and a large existing commercial infrastructure offers Sanger sequencing services. The resulting sequence information from our proposed amplicon area of the spike gene can be used to establish whether a patient has an infection with the current SARS-CoV-2 VoCs or any of the biologically important mutations in the spike gene. Finally, we provide a bioinformatics tool that enables automated basecalling, mapping and calling of mutations from Sanger trace files (.ab1). The results are reported in a table as well as in fastq format, available as a command line interface from https://github.com/kblin/covid-spike-classification or in a browser-based app running locally from https://ssi.biolib.com/app/covid-spike-classification/run.

**Figure 1.**
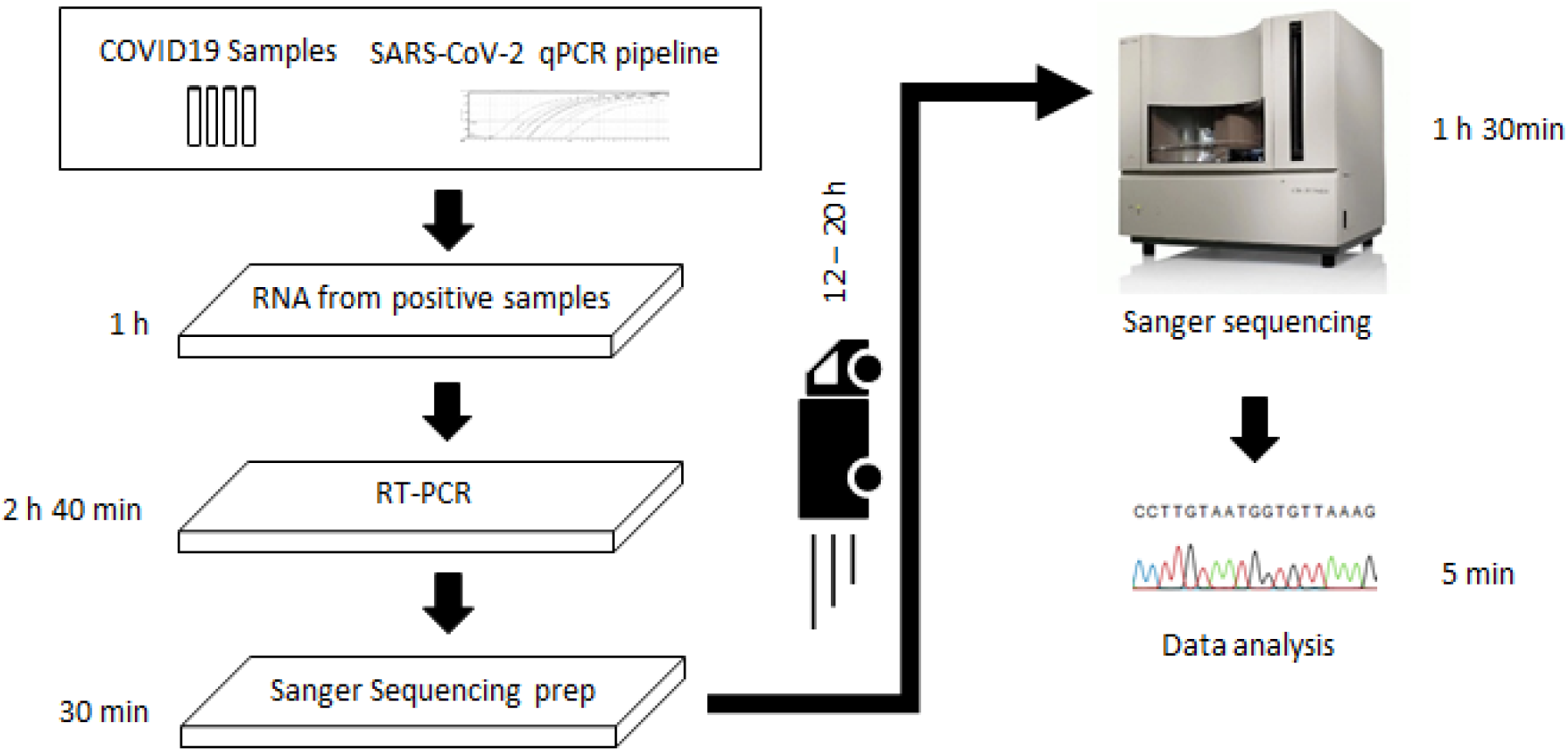
Workflow of the proposed Sanger sequence typing of SARS-CoV-2 variant mutations. Time estimates are based on a single 96 well PCR plate and no automation. The total hands on time is one hour 30 minutes, and only requires setting up an RT-PCR and transferring 1.5μl of the product to a new 96 well PCR plate, which is sent for Sanger sequencing at a commercial provider. The total cost per sample including plastware, enzymes, transport, and sequencing, is approx. 4 €. After the raw data is received, data analysis takes less than five minutes for 96 samples. If Sanger sequencing is available in-house, the time to result could be less than six hours.

## Methods

Purified RNA from the diagnostic RT-qPCR SARS-CoV-2 test facility at the Technical University of Denmark (DTU) was set up with two primers from the ARTIC Network SARS-CoV-2 v3 amplicon set for WGS v3 (https://github.com/artic-network/artic-ncov2019/blob/master/primer_schemes/nCoV-2019/V3/nCoV-2019.tsv) (Table 1). All assays were tested with RPC purified primers from TAG Copenhagen (Copenhagen, Denmark, http://tagc.dk). RNAdvance Viral Reagent Kit - Built on SPRI (Solid Phase Reversible Immobilization) bead-based technology (#C63510 Bechman Coulter, IN, USA), was used for the purification of input RNA. RT-PCR were set up in 20μl RT-PCR reactions using 10μl One Step PrimeScript III (Takara, Shimogyō-ku, Kyoto) used in the diagnostic SARS-CoV-2 test facility at DTU, 0.4μM forward primer, 0.4μM reverse primer, and 2-5μl purified template RNA per reaction (Table 1). The RT-PCR reaction was performed (after the initial test of annealing temperature) using the following program: reverse transcription for 5 min at 52 °C, then hot start polymerase activation at 95 °C for 10s, followed by 45 cycles of 95 °C for 5s, 58 °C for 30 s, and 72 °C for 1 min, followed by 5 min at 72 °C. This protocol is in line with the manufacturer’s instruction for a PCR product length of approx. 1kb. The resulting PCR products were analyzed using gel electrophoresis on a 1% agarose gel to confirm amplification (Suppl. Fig. 1). Then 10μl PCR product was purified with 0.8x volume of SPRI beads (KAPA Pure Beads, Roche, Basel, Switzerland), and both purified and unpurified PCR product was shipped to Eurofins Genomics (Eurofins, Luxembourg, Luxembourg) for Sanger sequencing using their Europe wide Mix2Seq Overnight Service. Samples were shipped at 15:00 one day and the Sanger sequencing results were received before noon the next day. We expect that any commercial Sanger sequencing provider can be used with the proposed protocol. For the proof of concept test with six samples, the recommended 5 ng/μl in 15 μl was shipped when possible, along with a final concentration of 1.2 μM left primer diluted in nuclease free water. For the larger scale test of 195 samples, 1.5 μl of unpurified PCR products were shipped. For RT-PCR product DNA concentration measurement, a Qubit 4 fluorometer was used with the dsDNA BR kit (Thermo Fisher Scientific, Waltham, Massachusetts, U.S.). Primer melting temperature was calculated using the Oligo Analyzer from IDT (https://eu.idtdna.com/calc/analyzer, Integrated DNA Technologies, Coralville, Iowa, USA). For analyzing the Sanger sequencing data, we set up a workflow consisting of: 1) basecalling ab1 files using Tracy^12^, followed by 2) mapping of fastq data to the NC_045512.2 SARS-CoV-2 sequence using Bowtie2^13^, and finally 3) mutation calling with Samtools^14^. Subsequently, the codons with known mutations are extracted and translated, and the amino acid calls are compared to a list of known mutations (currently N439K, L452R, Y453F, S477N, E484K, N501Y, A570D, Q613H, D614G, A626S, H655Y, Q677H, P681H, P681R, I692V, A701V, T716I). Information is primarily derived from covariant.org^15^), and reported as a csv table file, along with the basecalls in fastq format. The program is available from https://github.com/kblin/covid-spike-classification. Alternatively, Statens Serum Institut (SSI), the Danish government body for SARS-CoV-2 surveillance, has set up a self-contained web-app running the program without data upload at https://ssi.biolib.com/app/covid-spike-classification/run.

**Table 1.**
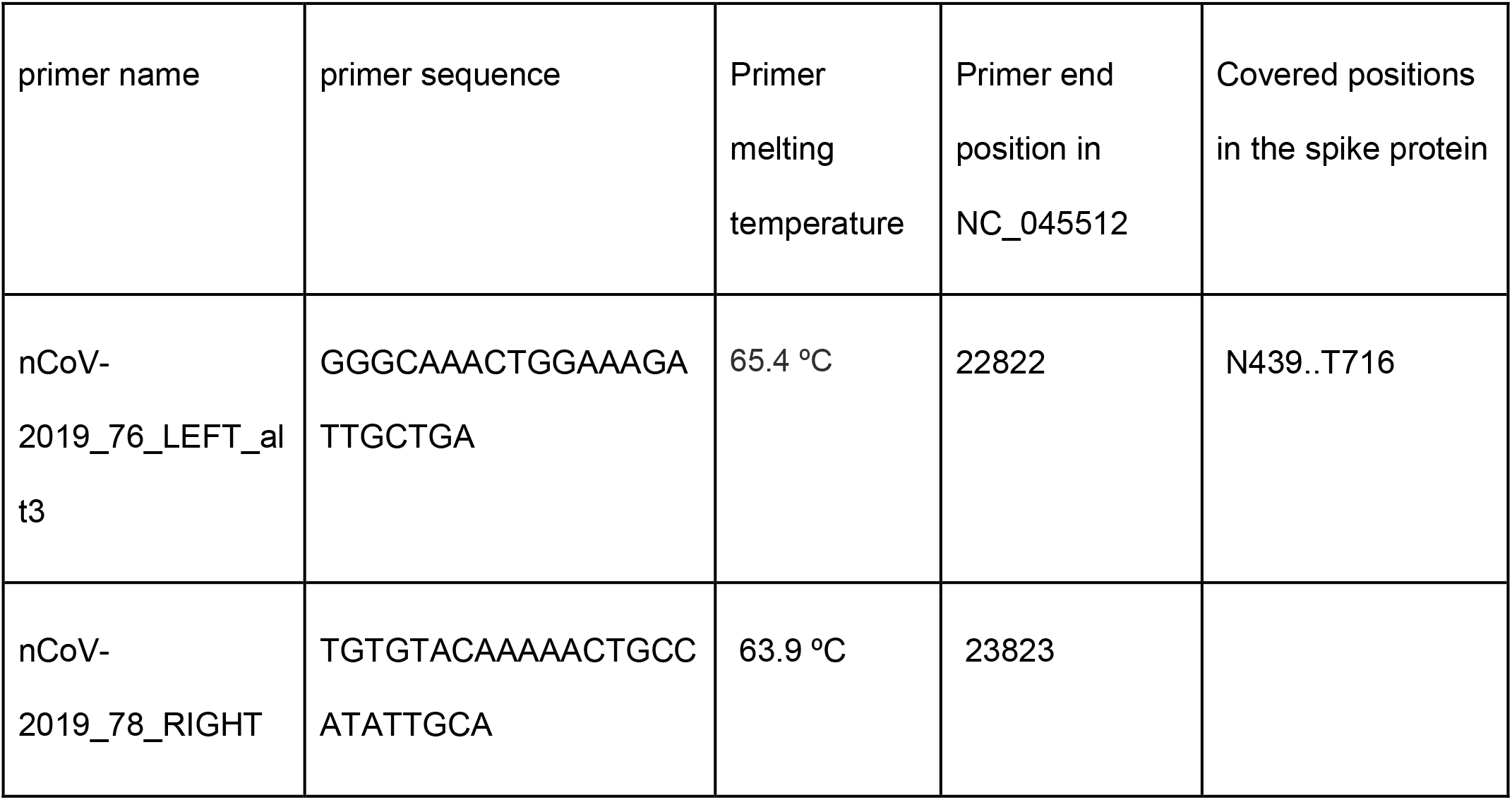
Primers suggested for typing SARS-CoV-2 VoCs. The Primer melting temperature is based on the use of Primescript III master mix, which has 2mM MgCl_2_.

The protocol for this method has been published at protocols.io at dx.doi.org/10.17504/protocols.io.bsbdnai6^16^.

The input RNA from the diagnostic SARS-CoV-2 RT-qPCR facility was obtained from oropharyngeal swabs with specimens placed immediately into a tube with 3M Guanidine Thiocyanate for instant lysis of viral particles. This ensures stability of RNA during transportation and during the process of transferring patient material from tube to plate. The RT-qPCR kit is demonstrated on PCR applications and has a Limit of Detection of 1 copy/µL for viral RNA (Centre for Diagnostics, DTU). RNA was purified in cooled racks and placed directly into the fridge prior to RT-qPCR reaction set-up. Following the reaction set up, the RNA was frozen at −20 C° prior to being thawed for sequencing within 1-2 days. For the diagnostic SARS-CoV-2 test, two separate primer sets and probes target the SARS-CoV-2 Nucleoprotein gene (N1 and N2), and one internal positive control primer set and probe which target the human RNAse P. A three-color multiplex probe system from Pentabase (Odense, Denmark) was used.

To estimate the profiling potential of the spike gene amplicon, SARS-CoV-2 strain data was collected from GISAID on 2021-01-29. Data was first pre-filtered by host “human” and minimum length > 25000. The data were then further filtered by excluding cases older than the first date at which all three VoCs appear (2020-12-04). This resulted in a dataset of 46,822 sequences. The VoCs B.1.1.7, B.1.351 and P.1 were assigned using Pangolin^17,18^ v2.1.7. To evaluate potential false-positive calls, a reference dataset of 5000 non-VoC samples was randomly selected from this dataset. To generate VoC reference data, the prefiltered data was randomly subsampled to a maximum of 1000 samples of each VoC. This led to downsampling of B.1.1.7 to 1000 and all P.1 (n = 40) and B.1.351 (n = 708) cases included. Sequences were assigned amino acid substitutions using Nextclade v0.11.2. To ensure amplicon dropout did not interfere with the evaluation, we removed samples which had >=50 consecutive bases missing within the amplicon region. This resulted in a final dataset of 878 B.1.1.7, 34 P.1, 516 B.1.351, and 4418 non-VoCs. We used the set of hallmark spike amino acid mutations that fall within the range of amino acid positions (position 419 to 759 on the spike protein), which are N439K, E484K, N501Y, A570D, H655Y, P681H, A701V, and T716I, along with the universal D614G mutation. Using the set of amino acid substitutions defined, we were able to compute all possible mutation combinations for each VoC within the amplicon window.

## Results & Discussion

Classification of SARS-CoV-2 genomes is often performed using Nextclade^19^ or Pangolin^18^ which uses several lineage defining mutations, but only a subset of mutations is needed as this allows profiling of incomplete genomes. Using 9 key amino-acid mutations included in a single 1001 bp amplicon in the spike protein gene (Table 2) from a total of 1428 VoC strains and 4418 non-VoC strains, we get an overview of the ratio of individual mutations for each of the B.1.1.7, B.1.351, P.1, and non-VoC strains (Accession numbers and references are stated in Suppl. Table 3). While nearly all strains shared the D614G mutation (99.8% of all strains), several mutations were exclusively or predominantly found in the VoC strains. The mutation N501Y was found in 99.9% of B.1.1.7, in 99.6% of B.1.351, and in 100% of P.1 of VoC classified strains (Table 2), while only present in 0.4% of non-VoC classified strains. Similarly, the mutation E484K was found in 99.2% of B.1.351 variants and in 100% of P.1 variants, but not in the included set of B.1.1.7 strains and in less than 0.1% of non-VoC strains. Several additional positions predominantly occur in VoC strains as shown in Table 2. We propose to use the Sanger sequence information in this amplicon to report and confirm individual mutations, which is sufficient to focus potentially limited contact tracing resources on patients who harbor VoCs, and it will provide enough information for surveillance of emerging variants.

**Table 2.** Key mutations in four SARS-CoV-2 variants and the potential to profile them based on a single amplicon in the spike protein. Note that even without the near universal D614G mutation, 100% of the VoC strains (n=1428) have one or more of the mutations, and less than 1% of non VoC strains would be wrongly interpreted as belonging to a VoC. The non-VoC lineage strains with e.g. E484K (four strains out of 4418) are important in their own right, as this mutation is thought to lower the current vaccine efficacy and this warrants extra contact tracing focus.

|  |  |  |  |  |
| --- | --- | --- | --- | --- |
|  | Pangolin lineage |  |  |  |
| Substitution | B.1.1.7 (n = 878) | B.1.351 (n=516) | P.1 (n=34) | non-VoC (n=4418) |
| N439K | 0% (0) | 0% (0) | 0% (0) | 7.4% (329) |
| E484K | 0% (0) | 99.2% (512) | 100% (34) | 0.1% (4) |
| N501Y | 99.9% (877) | 99.6% (514) | 100% (34) | 0.4% (16) |
| A570D | 100% (878) | 0% (0) | 0% (0) | 0% (0) |
| D614G | 99.9% (877) | 100% (516) | 100% (34) | 99.8% (4408) |
| H655Y | 0% (0) | 0% (0) | 100% (34) | 0.2% (11) |
| P681H | 99.8% (876) | 0.2% (1) | 0% (0) | 0.7% (31) |
| A701V | 0.1% (1) | 100% (516) | 0% (0) | 0.1% (4) |
| T716I | 99.8% (876) | 0% (0) | 0% (0) | 0.1% (6) |
| None of the<br>above<br>substitutions | 0% (0) | 0% (0) | 0% (0) | 0.2% (9) |

The proposed workflow for Sanger sequencing of SARS-CoV-2 positive samples uses a minimum of plastware and includes very few hands on steps (Figure 1). First, purified RNA from RT-qPCR positive samples are transferred to a 96 well plate. Then, the RT-PCR master mix is prepared in another 96 well plate and template is added. After RT-PCR, 1.5μl from each well is transferred to a PCR plate containing the Sanger sequencing primer and the plate is then ready for sequencing. The whole process requires only 3 pipette tips per sample, and approx. an hour and a half of hands on time (1 hour for RT-PCR preparation and 30 minutes for preparation of samples for sequencing), along with the two hour and 40 minutes run-time for the RT-PCR per 96 well PCR plate. The workflows proposed here are easy to automate. We estimate that the total price of these simple steps is 4€ per sample in material cost including plastware, enzymes, buffers, shipping, and sequencing, making the workflow cost efficient compared to WGS or multi-target RT-qPCR typing approaches. If Sanger sequencing can be performed in-house the time to result can be less than six hours from positive COVID-19 sample to variant call, which is faster than any high throughput WGS based analysis, with less computational workload and data storage.

To evaluate the suitability of the Sanger sequencing method for mutation calling in the RT-PCR product, we initially used six SARS-CoV-2 positive RNA samples from the large scale testing facility at DTU. We shipped both SPRI purified and unpurified RT-PCR products for Sanger sequencing (Table 1, Suppl Figure 1). In this way, we got two Sanger reads from each of the 6 samples, which we used to verify that the mutations are called identically regardless of purification status. Suppl. Table 1 shows an overview of the 12 Sanger reads. We tested a custom made primer set with similar results (Suppl. Table 1). A single sample had mutation calls identical to B.1.1.7 variants in all four reads, as it had the key mutations N501Y, A570D, and P681H in all four Sanger reads. This successful proof of concept study shows that the single ARTIC primer set can be used for typing current SARS-CoV-2 VoC key defining mutations and that unpurified RT-PCR product can be used, significantly reducing the workload involved in mutation typing. Following RT-PCR and shipping of a larger batch of 195 samples for Sanger sequencing, we received the results within 20 hours, which is routine delivery time for most large commercial providers. In Figure 2, the 195 samples have been binned according to the diagnostic RT-qPCR Ct value for both the N1 and N2. Note that the initial seven cycles are not fluorescence recorded and therefore do not count towards the final Ct value in the RT-qPCR set up used at the DTU testing facility. Overall, the success rate of Sanger sequencing of the RT-qPCR positive samples was 85.1% (166/195 Sanger reads mapped to the correct position of the SARS-CoV-2 spike gene). If failed samples were rerun or duplicate PCR reactions implemented, we expect that a slightly higher fraction of samples would yield mutation calls. The fraction of samples with a mappable result from Sanger sequencing is stable at approx. 90% below Ct 30 (Figure 2, note the seven initial unrecorded cycles). For samples where the Sanger read successfully mapped, a total of 15 positions out of 1,494 (166 mapped samples and 9 investigated targets) were not assigned a value (1.0% of positions, see Suppl. Table 2), mainly from the ends of the sequence. As expected, the “No Template Control” did not yield a result. Five of the 166 sequences had the combination of the N501Y, A570D, P681H, and T716I mutations, which strongly suggests infection with the B.1.1.7 lineage (Table 2, Suppl. Table 2).

**Figure 2.**
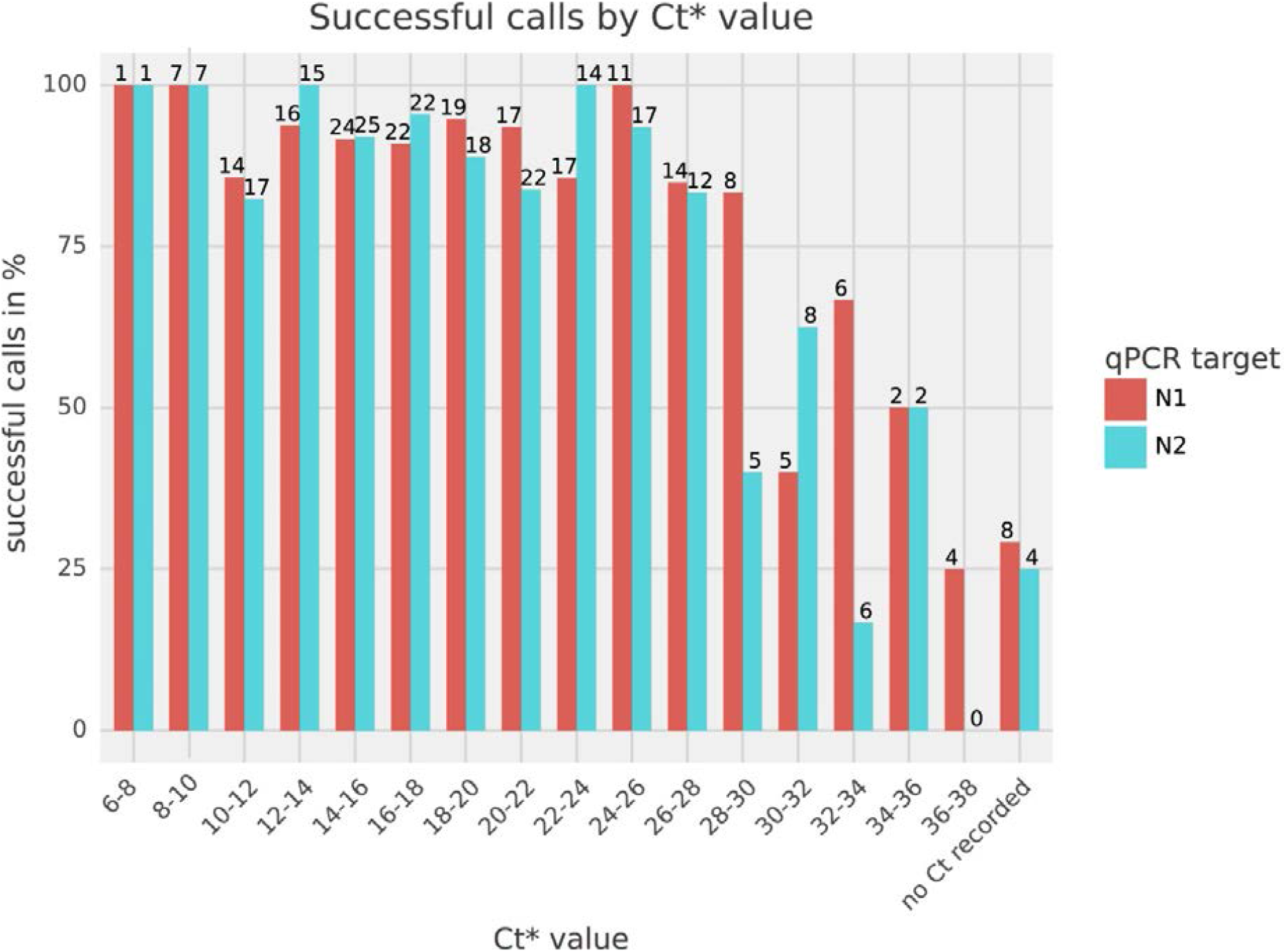
Sensitivity of Sanger sequencing of the proposed amplicon compared to the diagnostic RT-qPCR test (Ct values, n=195). The numbers above each bar refer to the number of samples in that bin. Overall, 85.1% of samples were of sufficient quality for mapping. The difference between N1 and N2 reflects that the Ct for the two RT-qPCR targets can be different, and sometimes one target is not recorded (n=8 for N1 and n=4 for N2). The NTC is not depicted, as it did not produce a Sanger read that mapped to the reference genome. *: Ct values stem from a 2-step PCR where the first 7 cycles are not fluorescence registered.

The enzymes we used in the proposed RT-PCR set up are identical to those used in the COVID-19 diagnostic test facility at DTU, where up to 10,000 patient laryngeal swab samples are processed per day. Because the enzymes are identical, the supply chain is simplified. It is likely that any RT-PCR enzyme mix will perform well with our setup. Identifying emerging VoCs requires large-scale tracking of the complete SARS-CoV-2 genome, and we do not recommend substituting the WGS efforts with Sanger sequencing of a single area of the spike gene. However, spike-gene Sanger sequencing of all SARS-CoV-2 positive samples can enable rapid mutation typing and association to current variants and mutations of concern, and potentially the proposed amplicon can cover mutations arising in the future. In February 2021, there are currently two additional lineages of note, B.1.525 and A.23.1, that contain key defining mutations within the amplicon sequenced. RT-qPCR with primers and probes specific for VoC mutations is being used to monitor outbreaks, and while this method is arguably faster than Sanger sequencing, there are several drawbacks. For instance, RT-qPCR assays usually use several primers and probes, rendering it more expensive than the proposed Sanger sequencing based method. qPCR assays can only investigate a few mutations in a sample, where Sanger sequencing gives the sequence over a 1 kb part of the spike gene of SARS-CoV-2. In this way, new VoCs are likely to be detected without change to the Sanger sequencing setup, whereas a VoC specific RT-qPCR setup would likely require extensive testing of a new primer probe combination each time a new VoC is classified.

## Conclusion

We show that typing of current SARS-CoV-2 VoC specific mutations can be performed by Sanger sequencing of an RT-PCR product from the spike gene using a single primer set and existing commercial sequencing infrastructure in a cheap and fast way. This requires no other enzymes or equipment besides those already in daily use at any SARS-CoV-2 RT-qPCR testing facility. Our protocol has the potential to change how SARS-CoV-2 VoC mutations are typed globally, as it is the cheapest and simplest way proposed yet to yield sequencing information from SARS-CoV-2 positive samples. We achieved mutation calls from 85.1% of all samples, some of which only recorded one of two targets in the RT-qPCR diagnostic SARS-CoV-2 test. We do not suggest abandoning the use of WGS, as it is important for tracking emerging variants. The proposed workflow is currently being used routinely to type all positive samples at the High Throughput SARS-CoV-2 RT-qPCR testing facility at DTU.

## Statement of data use

All samples are pseudoanonymized before arrival at the DTU facility, and once more before shipment of samples to Eurofins genomics. For this study, Sanger reads were anonymized.

## Data Availability

Raw data will be made public after peer review.

## Acknowledgements

We thank Jeppe Hallgren and Jørn Emborg from biolib for setting up the self-contained web-app and Kim Ng and Thor Bech Johannesen for coordination assistance from Statens Serum Institut. We thank Benni Winding Hansen for equipment access. We thank the NGS lab at The Novo Nordisk Center for Biosustainability for allowing parts of the work to be performed in their facilities. We thank Søren Karst for valuable discussions. This work was supported by the Poul Due Jensen Fonden (Grundfos Fonden, Corona-Danica). T.S.J. and K.B. received support by The Novo Nordisk Foundation through the grants NNF-IIMENA (NNF16OC0021746) and NNF CFB (NNF20CC0035580). We are gratefully to researchers, clinicians, and public health authorities for making SARS-CoV-2 sequence data available through GISAID (supplementary table 3).

---

**Suppl. Fig. 1.**
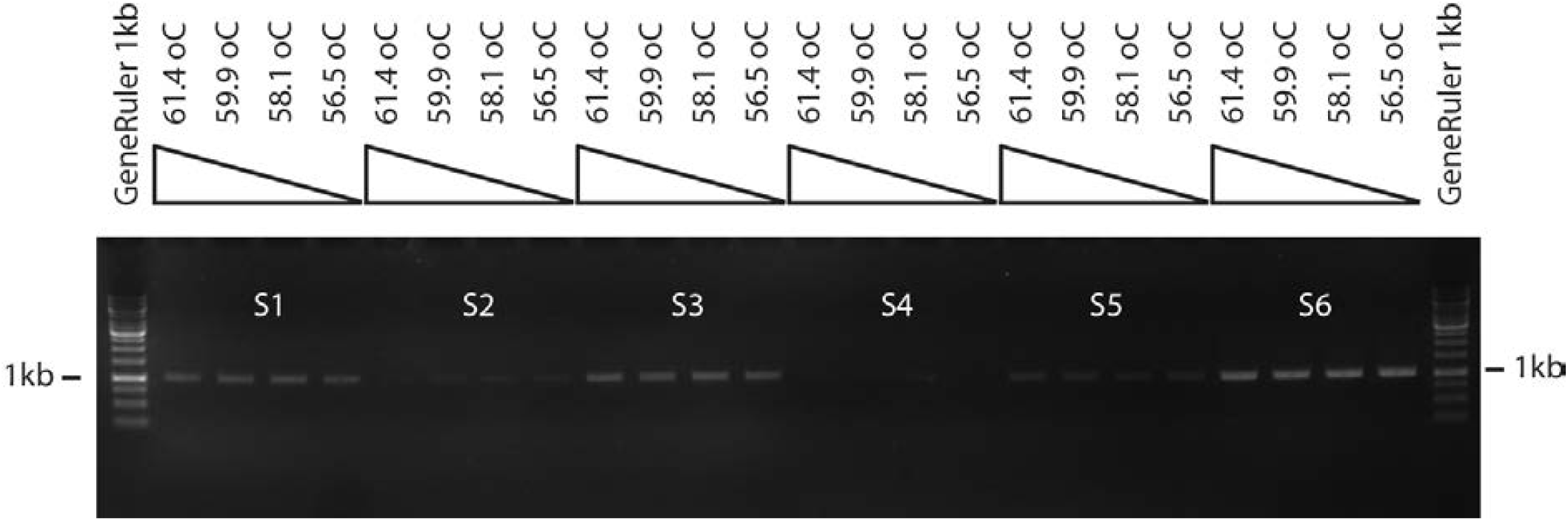
1% Agarose gel electrophorese of primer test. Each well has 1.5μl RT-PCR product regardless of DNA concentration.

Suppl. Fig. 1: PCR primer annealing temperature test

Suppl. Table 1: Proof of concept DNA concentration of 24 samples, Ct values and mutation calls

Suppl. Table. 2: Ct values of 195 samples and mutation calls

Suppl. Table. 3: GISAID references

## Notes

### Competing Interest Statement

The authors have declared no competing interest.

### Author Declarations

A waiver of ethical approval was received from the regional Copenhagen ethics committee. The decision to waive ethics approval was made by Cand.jur Jakob Hjelvang Lemming,, Region Hovedstaden, Center for Regional Udvikling, Sundhedsforskning og Innovation, De Videnskabsetiske Komiteer for Region Hovedstaden, Kongens Vaenge 2, 3400 Hilleroed, Denmark

